# An evidence synthesis approach for combining different data sources illustrated using entomological efficacy of insecticides for indoor residual spraying

**DOI:** 10.1101/2021.05.11.21257015

**Authors:** Nathan Green, Fiacre Agossa, Boulais Yovogan, Richard Oxborough, Jovin Kitau, Pie Müller, Edi Constant, Mark Rowland, Emile FS Tchacaya, Koudou G Benjamin, Thomas S Churcher, Michael Betancourt, Ellie Sherrard-Smith

**Author notes:** **Corresponding author:** Nathan Green.

## Abstract

**Background:** Prospective malaria public health interventions are initially tested for entomological impact using standardised experimental hut trials. In some cases, data are collated as aggregated counts of potential outcomes from mosquito feeding attempts given the presence of an insecticidal intervention. Comprehensive data i.e. full breakdowns of probable outcomes of mosquito feeding attempts, are more rarely available. Bayesian evidence synthesis is a framework that explicitly combines data sources to enable the joint estimation of parameters and their uncertainties. The aggregated and comprehensive data can be combined using an evidence synthesis approach to enhance our inference about the potential impact of vector control products across different settings over time.

**Methods:** Aggregated and comprehensive data from a meta-analysis of the impact of Actellic® 300CS (Syngenta), an indoor residual spray (IRS) product, used on wall surfaces to kill mosquitoes and reduce malaria transmission, were analysed using a series of statistical models to understand the benefits and limitations of each.

**Results:** Many more data are available in aggregated format (*N* = 23 datasets, 5 studies) relative to comprehensive format (*N* = 3 datasets, 2 studies). The evidence synthesis model was most robust at predicting the probability of mosquitoes dying or surviving and blood-feeding. Generating odds ratios from the correlated Bernoulli random sample indicates that when mortality and blood-feeding are positively correlated, as exhibited in our data, the number of successfully fed mosquitoes will be under-estimated. Analysis of either dataset alone is problematic because aggregated data require an assumption of independence and there are few and variable data in the comprehensive format.

**Conclusions:** We developed an approach to combine sources from trials to maximise the inference that can be made from such data. Bayesian evidence synthesis enables inference from multiple datasets simultaneously to give a more informative result and highlight conflicts between sources. Advantages and limitations of these models are discussed.

**Key messages:**

- Statistical models are developed to best infer key mosquito IRS statistics from disparate data sources.
- Naïve methods, previously adopted, of combining different data sources can create bias and must be explicitly considered.
- Mosquitoes are highly likely to feed in experimental hut trials using indoor residual spraying of insecticides so the assumption of independence in the probability of mosquitoes dying is reflective of trial data.
- We provide code for practitioners to implement this approach themselves in both BUGS and Stan languages running from R.

## Introduction

Interventions that shorten the mean lifespan of a mosquito and interrupt biting cycles are integral to the control of malaria infections across Africa^1^. The entomological effect of the indoor residual spraying of insecticide (IRS) and insecticide-treated nets are tested using experimental hut trials as part of the product validation process and before World Health Organization (WHO) recommendations can be considered and made^2^. These trials apply IRS to huts and then the temporal impact of the IRS is tracked using a human study participants who stays overnight to act as bait for blood-seeking mosquitoes. Over the course of the malaria transmission season, when mosquitos are present, the outcomes of mosquito mortality and feeding attempts are observed and compared to those incurred for a study participant who stays overnight in an unsprayed hut. On entering an IRS treated hut, a mosquito may: i) blood-feed on human-baits; ii) be killed by the IRS chemical compound, or; iii) exit into window or veranda traps.

These hut trial data are often published in aggregated format as the key metrics outlined by WHO (induced mortality, blood-feeding inhibition and deterrence)^2^ do not need data to be disaggregated. The effects of interventions must be replicated in multiple settings that have different ecological characteristics to better understand the overall protection that IRS – or other vector control – can afford. Systematic reviews are increasingly used to assess ecological trends in these combined data and summarise evidence to help evaluate interventions^3–5^.

Compiling aggregated data (AD) such as experimental hut studies can cause complications for meta-analyses that use the data for slightly different purposes because, among other reasons: i) AD can be presented in inconsistent ways by summarising results^6^ making data hard to harmonize across different trials^7^; ii) AD may not fully account for individual-level characteristics leading to ecological bias^8,9^; iii) if large numbers of large trials are available, meta-regression analyses of AD may prove statistically powerful but with few or smaller trials AD may miss clinically significant treatment level effects^10^, and; iv) within study variability may be missed^11^. However, there are often few data sets available for any intervention tested and, historically, only a subset of these trials may provide individual-level data (ILD), which could alleviate some of these issues.

In the experimental hut data testing IRS products, a specific challenge arises from aggregating data because no distinction is made as to whether mosquitoes have blood-fed and survived or blood-fed and been killed. This is an important epidemiological distinction because those mosquitoes that blood-feed and survive may go on to oviposit or transmit malaria parasites onward. Our recent assessment of IRS products made the simplifying assumption that mosquitoes were equally likely to have blood-fed and survived or blood-fed and died on entering a sprayed hut^3^. In this paper, we use systematically collated data from Sherrard-Smith *et al*.^3^ on the IRS product Actellic 300®CS (Syngenta) as an example dataset to explore different models that aim to infer how the impact of the vector control product on mosquitoes changes over time using both AD and ILD. Within these data we have 5 studies presenting aggregated data (23 time series), of which 2 studies also present comprehensive (ILD) data (3 time series). Careful consideration is required particularly for any analysis where data are aggregated in different ways across trials, and where the comprehensive data are only part of the total available data. Ideally, we want to ensure that any inference using AD is in agreement with inference afforded by the comprehensive data. Bayesian statistical methodologies provide a natural paradigm to analyse evidence from multiple sources in different formats^11–16^. Bayesian evidence synthesis is a framework that explicitly combines data sources enabling joint estimation of parameters and their uncertainties^17^. We compare predictions of each statistical model presented to those estimated by analysing the subset of either aggregated data or comprehensive data individually. We demonstrate the advantage of inferring from both data sources.

## Methods

### Data

Actellic® 300CS is an IRS product consisting of the organophosphate insecticide pirimiphos-methyl that is widely used for IRS campaigns across the African continent since WHO recommendation in 2013^18^. The product was evaluated using West African experimental huts in Benin, Côte d’Ivoire and Tanzania across 5 studies^19–22^ [one unpublished]. Twenty-three datasets from these studies had aggregated data reporting the total number of mosquitoes that entered sprayed huts, and the total number killed, blood-fed or exited without the distinction for comprehensive assessment for at least 3 time points (Supplementary Data 1, dataset 1). Comprehensive data were available from three of these datasets (data available upon request) (data resources are summarised in Table 1). At least 3 repeated measures through time were made for each study ranging from 6 to 12 months since the insecticide was first deployed.

**Table 1:**
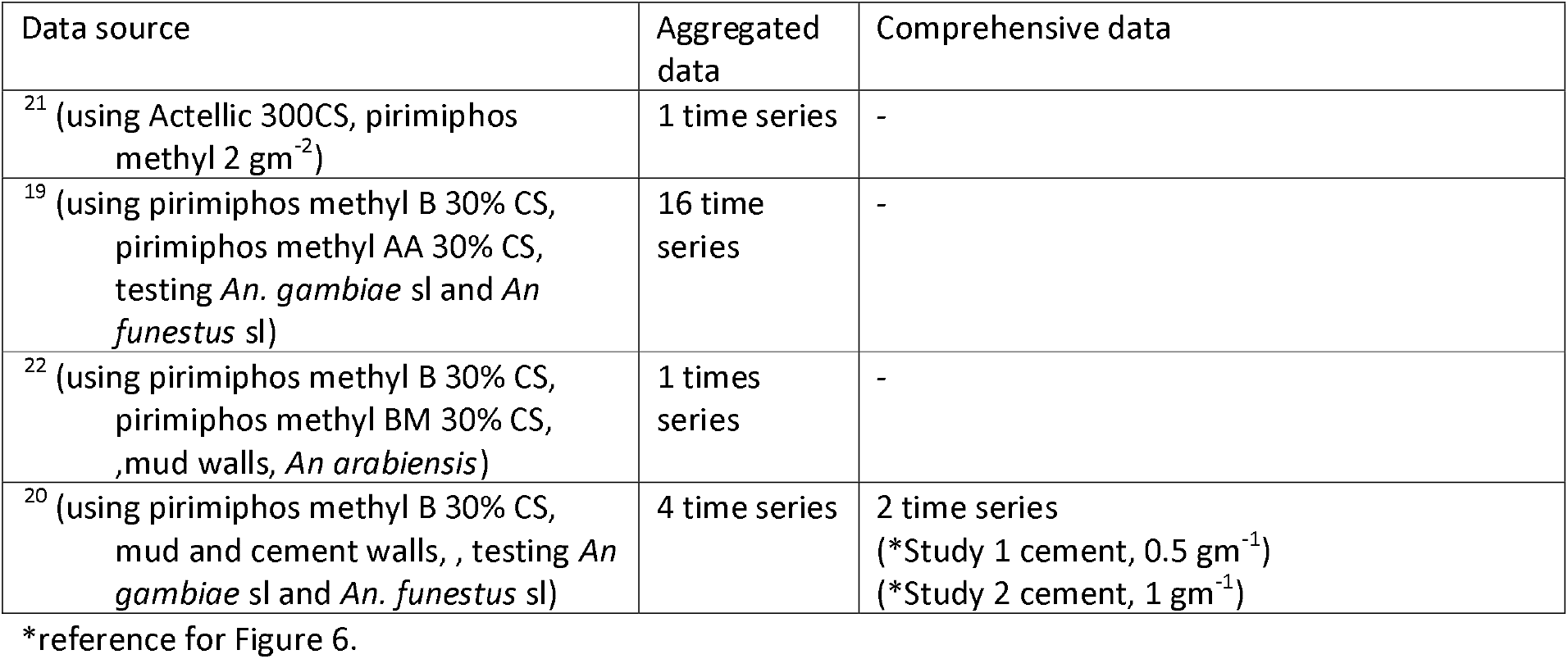
A list of the studies included and which models are informed by each dataset. Where indicated, the data are provided in full, in Supplementary Data file 1.

Our aim is to determine the probability that a mosquito is either killed or blood-fed and surviving after attempting to feed in an experimental hut sprayed with insecticide. We determine how this association changes over time and how probabilities differ between mosquito species.

A contingency table to demonstrate the different aggregation of these data from the available sources is shown in Figure 1.

**Figure 1:**
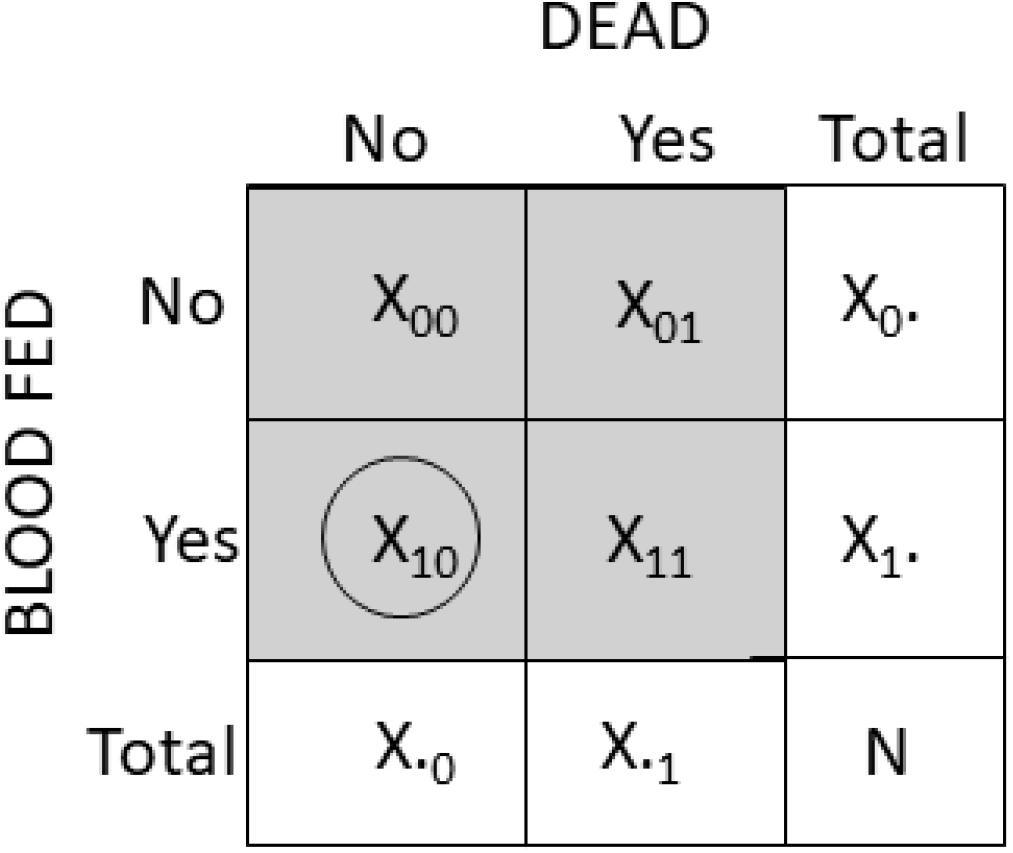
Contingency tables of trial data: The number of mosquitoes that are blood-fed (X_1_.) and the number of mosquitoes that are killed (X._1_) are known for both data sources but, for the aggregated data we do not know directly the number of mosquitoes that are both alive and blood-fed (X_10_) (circled). We have the full complement of data from the comprehensive source.

### Statistical methods

We built three statistical models that were fitted in a Bayesian framework using the probabilistic programming software OpenBUGS (release 3.2.3)^23^, accessed through R (version 3.6.1)^24^. Data are provided in Supplementary data file 1, model code are provided in Supplementary Material 1. We provide equivalent code for the same process to be conducted in RStan (version 2.21.1)^25^ in Supplementary Material 1.

The first model (demonstrated schematically in Figure 2a, Model 2a) uses the aggregated data only and estimates the proportion of mosquitoes that are killed and those that are blood-fed before inferring from the product of these probabilities, those that are surviving and blood-feeding, using the assumption that a mosquito is equally likely to be blood-fed whether killed or not.

**Figure 2:**
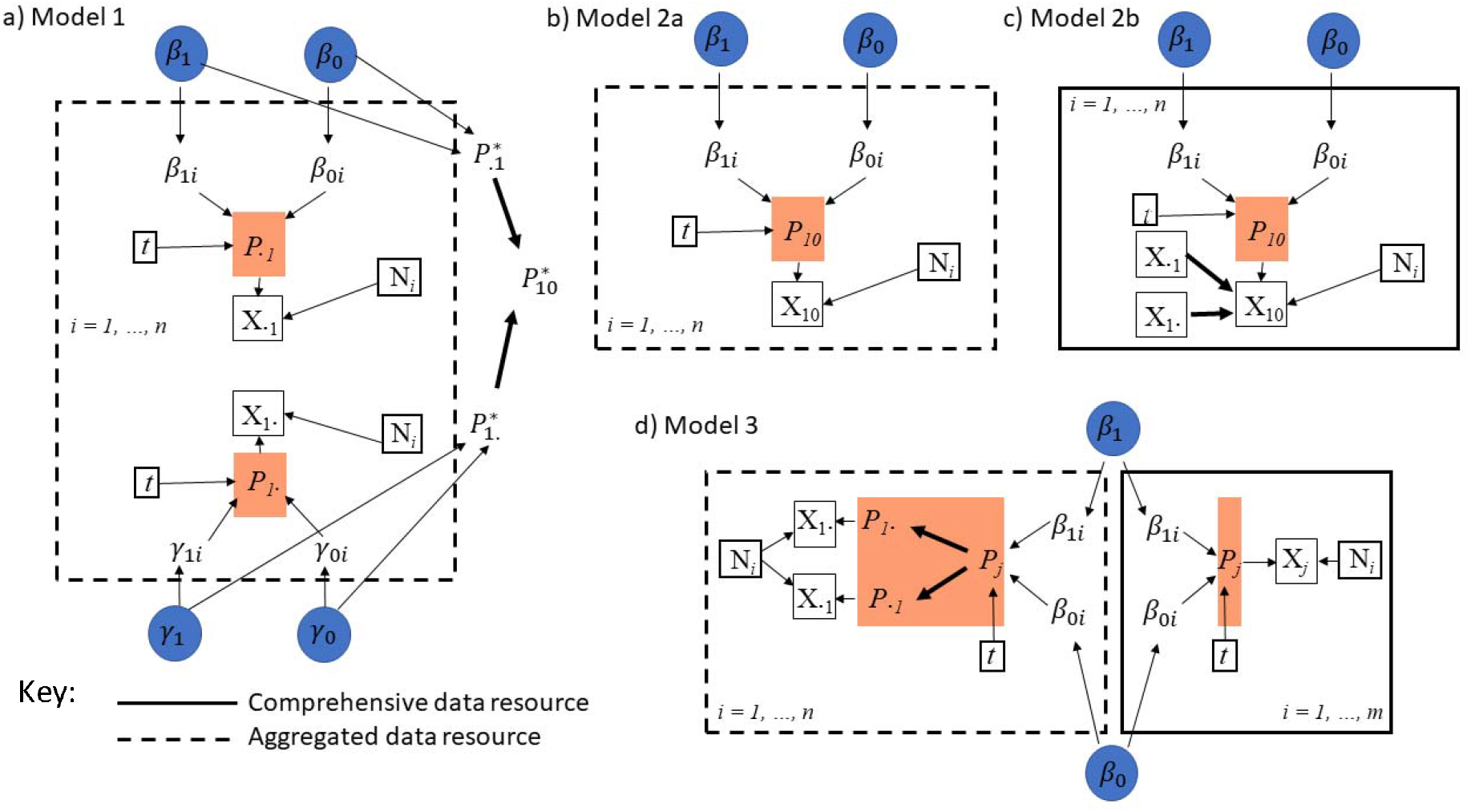
Directed acyclic graphs (DAGs) for models to assess aggregated (a, b), comprehensive (c) or both aggregated and comprehensive (*d*) data. (a) Mosquitoes are counted as killed with no information on whether they are also blood fed (*X*_*1*_.) or unfed without information that mosquitoes are also killed (*X*_.1_). We can infer marginal global probabilities (shown in orange boxes) then (assuming independence), after estimating the probability of either outcome (*P*_.1_* and *P*_1_.*), infer the probability that mosquitoes are both alive and blood fed (*P*_01_*). b) Alternatively, we can adjust the data using the same assumption of independence prior to fitting the model and fit a logistic binomial model to the adjusted data. c) The same model structure (Model 2) can be fitted to the comprehensive data. d) Using evidence synthesis, we can learn from the comprehensive data (*N* = 3 datasets) to infer probabilities that are supported by the aggregated data (*N* = 23 datasets). In each DAG, *j* represents the 4 potential outcomes from a feeding attempt, (*P*_*01*_ *P*_*00*_ *P*_11_ or *P*_10_) for each dataset *i* over time *t*.

The second model adjusts the aggregated data directly prior to fitting the model and then fits a logistic binomial to estimate the respective probabilities from the adjusted data. This is an approach used previously^3^ on aggregated data (Figure 2b, Model 2a). This model structure is also fitted to the comprehensive data where no prior assumptions are required (Figure 2c, Model 2b) to estimate the probabilities from the comprehensive data set.

The third model (Figure 2d, Model 3), a Bayesian evidence synthesis model, is constructed similarly but distinct from previous methods^16,17,26^. This model combines the data sources probabilistically to incorporate the inferences that can be made from the comprehensive data benefited by the additional aggregated data source.

### Model 1

The largest number of available trials provide AD. Model 1 (Figure 2a) (and Model 2, Figure 2b) disregards the smaller subset of richer ILD. These trials do not record which mosquitoes had jointly survived and blood-fed. Posterior distributions are estimated for the marginal probabilities of mosquitoes that are blood-fed (*P*^*f*^) or mosquitoes that have been killed (*P*^*d*^) (Figure 1).

A time-dependent logistic function is fitted to IRS impact on mosquito mortality and mosquito blood-feeding (*t*, in days).

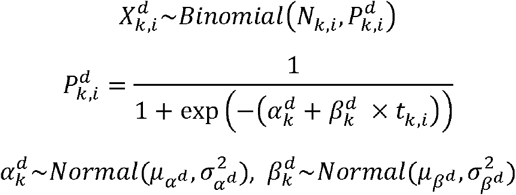

Superscript denotes whether that mosquitoes will die (*d*). The subscripts denote the trial identifier *k* and the time point at which measurements are taken *i*. We use equivalent formulae for the fed model where blood-fed (*f*) replaces mosquitoes that die (*d*). Parameters *α* and *β* determine the shape of the binomial relationship and have normally distributed priors with mean *µ* and variance *σ*^*2*^. The raw data are indicated by *X*, the count of mosquitoes that were killed (or blood-fed), and *N*, the total count of mosquitoes.

Given the absence of data on the mosquitoes that have survived and blood-fed, we can instead infer this probability by assuming independence, combine the marginal probabilities to estimate the joint probability that mosquitoes are both dead and blood-fed:

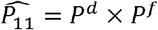

Or those that survived and fed:

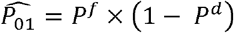

In other words, we assume that mosquitoes are equally likely to have been killed and blood-fed or to have survived and blood-fed (Figure 3a and 3b).

**Figure 3:**
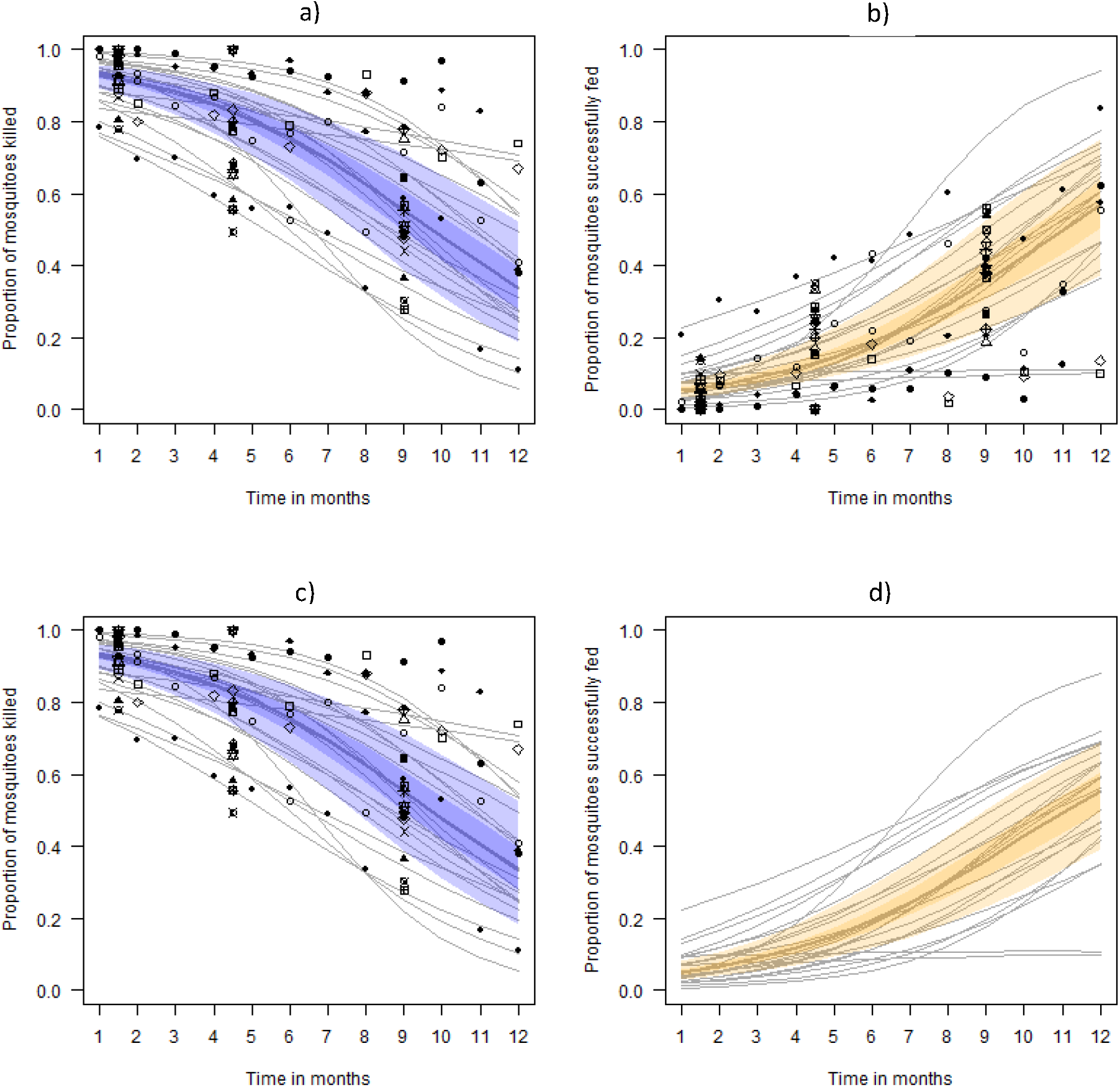
Model predictions for the best-estimate (median posterior predictive value) across all study datasets, 50% (darker shaded region) and 95% (lighter shaded region) credible intervals (CrI) from the respective fits to aggregated data. Data for each fit are overlaid onto the figure to demonstrate the suitability of the time-dependent functions. In each of the fits, the individual study predictions are overlaid on the figures by thin grey lines and noted by matching symbol type for each timeseries. Mortality data are identical for Models 1 (a) and 2 (c). The blood-fed data are only available in aggregated format and are either probabilistically (b) or empirically (d) estimated by the respective models. *Anopheles gambiae* s.l. (circles), *An. funestus* s.l. (squares) and *An. arabiensis* (triangles) mosquitoes are noted. The data from the comprehensive source are only used in aggregated format and shown for Benin (dotted line), and Côte d’Ivoire (dashed line).

### Model 2

We contrast this with a previous approach^3^, where, before fitting the time series data, the number of mosquitoes that were blood feeding *(X*_*1*_.) is adjusted by assuming that blood-feeding does not increase the probability of being killed. The blood-feeding and surviving mosquitoes are assumed to be:

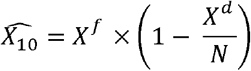

A time-dependent logistic function is fitted to these adjusted data on IRS impact for mosquito survival and successful blood-feeding (*t*, in days) (Figure 3c and 3d):

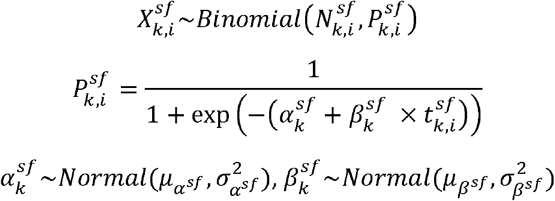

This model is also fitted to the comprehensive data only to estimate the four different potential outcomes from a mosquito feeding attempt (Figure 2c, Figure 4a and 4b) shown in grey boxes in the contingency table (Figure 1).

**Figure 4:**
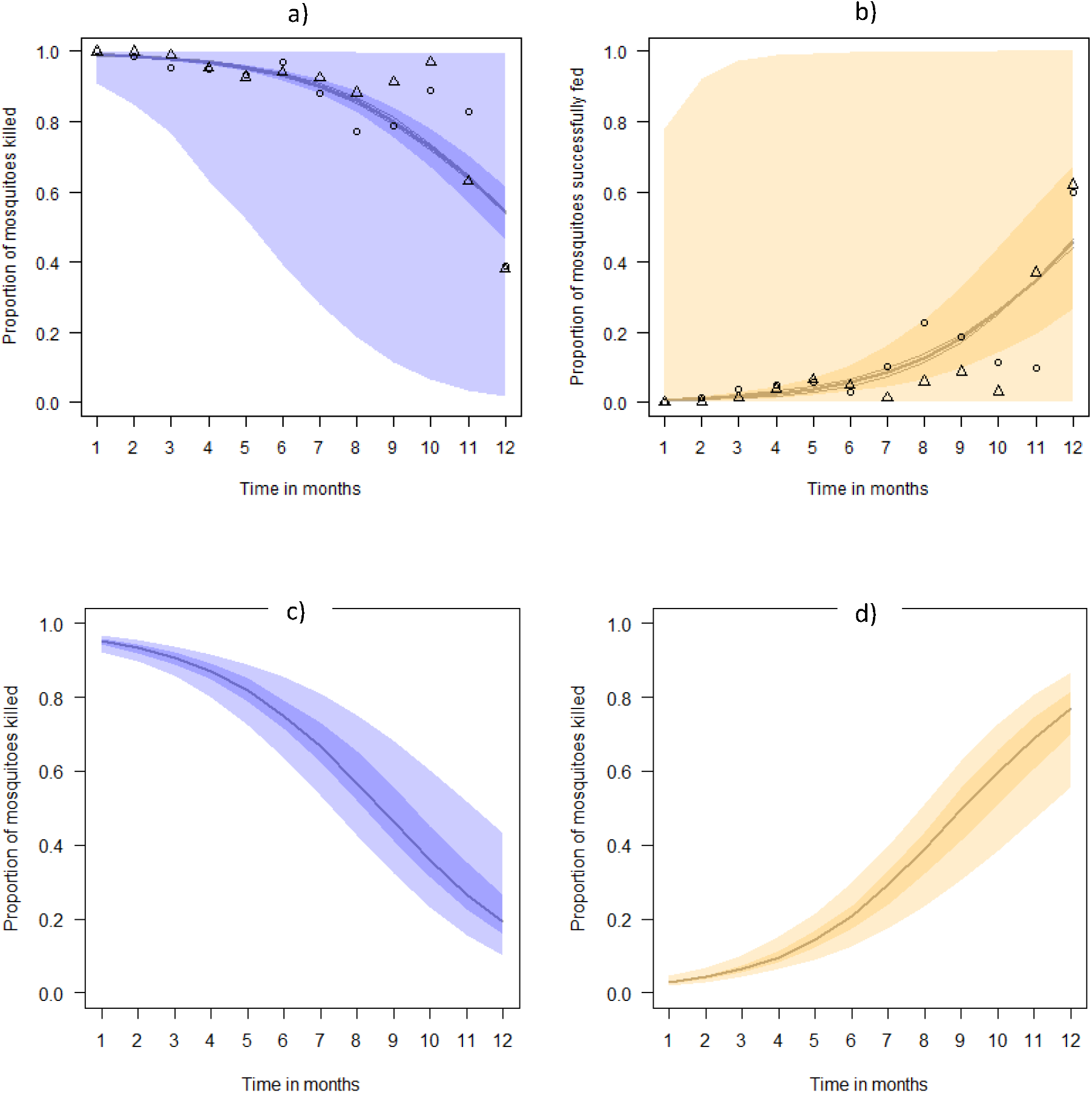
Model predictions for the best-estimate (median posterior predictive value) across all study datasets, 50% (darker shaded region) and 95% (lighter shaded region) credible intervals (CrI) from the respective fits to the comprehensive data. Model 2 (a and b) is also fitted to the comprehensive data for which there are fewer data (N = 3 datasets). Model 3 (c and d) is informed by both aggregated and comprehensive data to predict the probability that a mosquito will be killed or will survive and blood-feed. *Anopheles gambiae* s.l. (circles), *An. funestus* s.l. (squares) and *An. arabiensis* (triangles) mosquitoes are noted. Symbol shapes correspond to the different study data fitted (overlaid onto a and b) (Benin dotted line, Côte d’Ivoire dashed line).

### Model 3

Model 3 presents the Bayesian evidence synthesis that allows inference of the parameter estimates for each potential mosquito outcome through combining both data sources.

AD are assumed to fit binomial distributions such that:

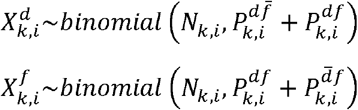

A logistic function is used to describe the time-dependent data:

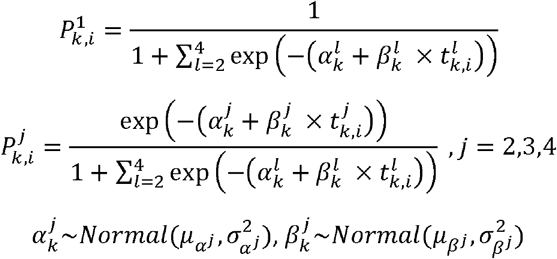

The superscript *j* represents the possible outcomes noted in the contingency table (Figure 1).

The comprehensive data are modelled similarly but using a multinomial distribution for the 4 categories and the logistic link function.

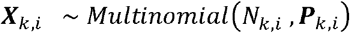

Where *X= (X*_*00*,_ *X*_*10*,_ *X*_*01*,_ *X*_*11*_) and 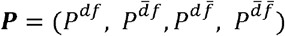.

The hyper-parameters to generate the posterior predictions for each distribution are linked across the two datasets such that they are exchangeable, i.e. for *k* consisting of both ILD and AD trials

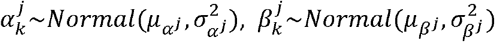

To understand better how the variability in data sources effects predictive ability of the evidence synthesis model, we generated correlated Bernoulli random samples where the odds ratio for each source was either matched, i.e. the predicted mosquitoes surviving and blood-feeding are reflected in both data sources, or mis-matched, i.e. the aggregated source over- or under-estimates the proportion of mosquitoes that survive and blood-feed (Figure 5).

**Figure 5:**
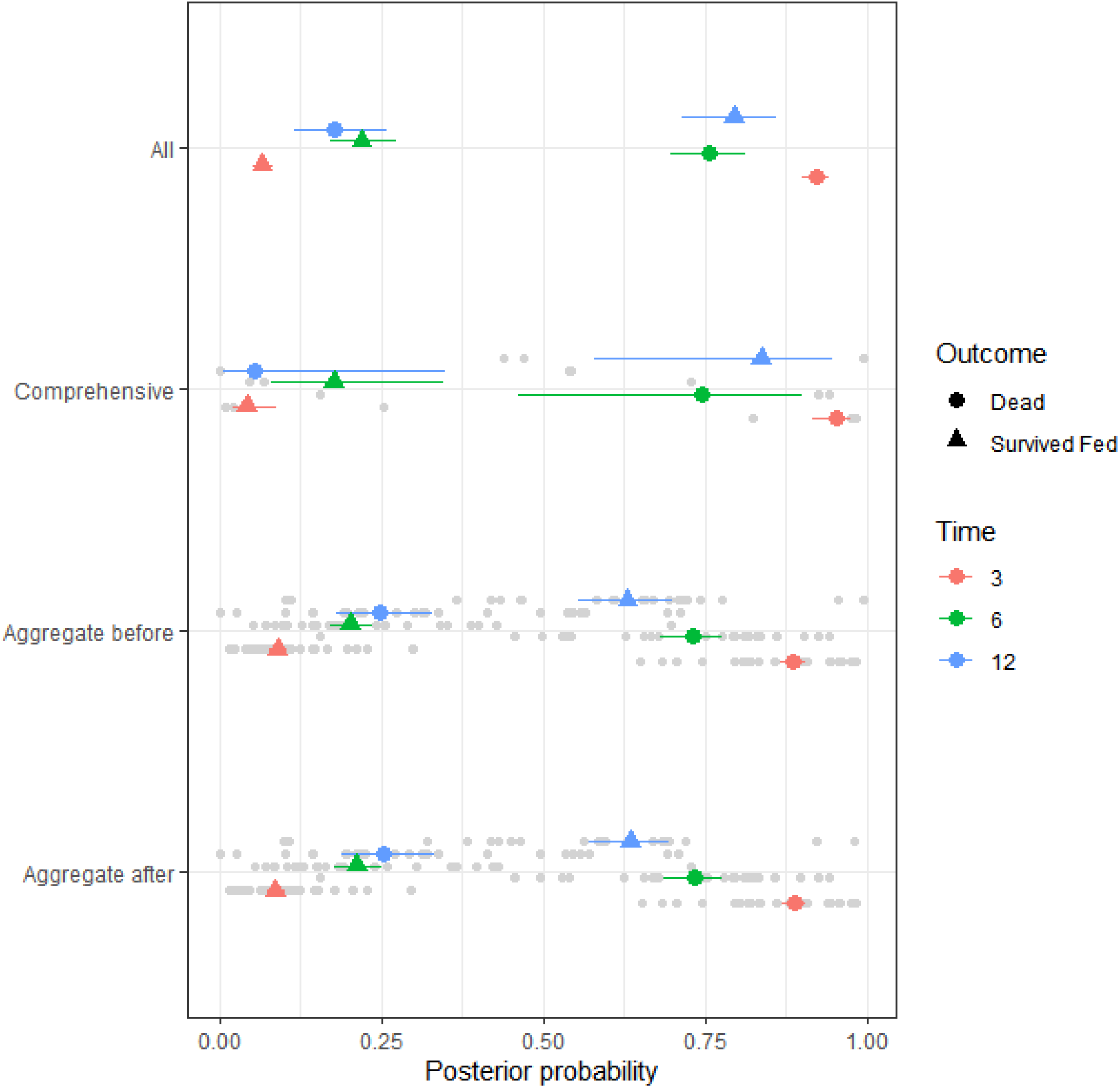
A forest plot of posterior predictions for all models, showing median, 25% and 75% percentiles for dead and survived fed probabilities across all study datasets at 3, 6 and 12 months. Individual study predictions are shown with grey points.

**Figure 6:**
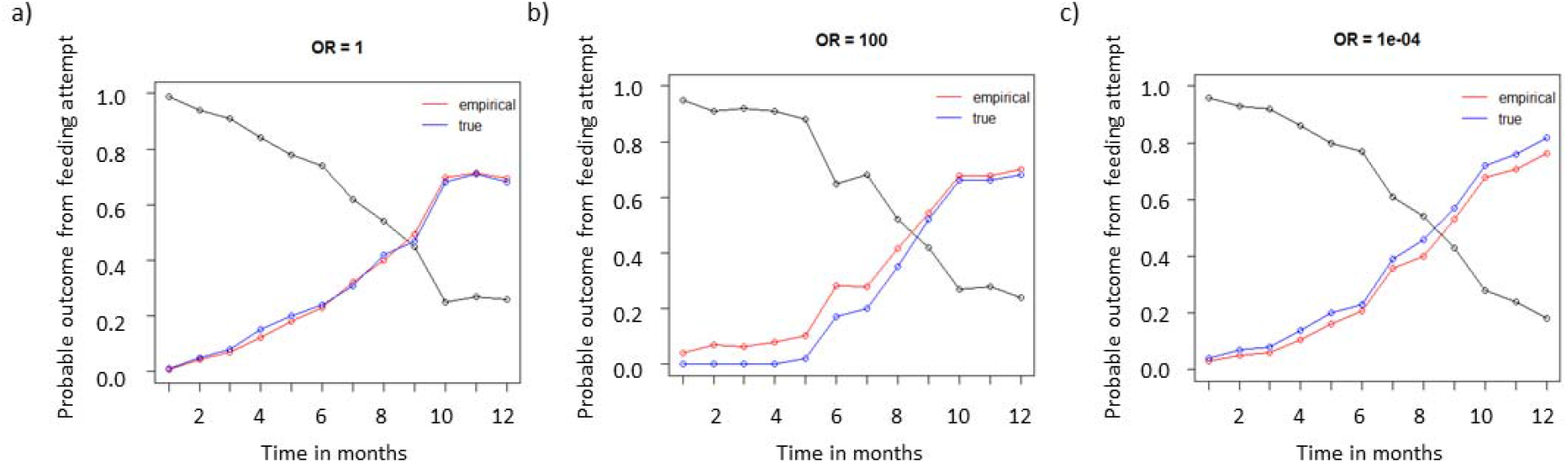
The bias of the evidence synthesis approach given different odds ratios (OR) between simulated aggregated and comprehensive data sets. Weighted proportions survived and fed over time for the true (blue) and empirical (red) estimates. The empirical are the product of the marginal probabilities thus assuming independence. a) An odds ratio of 1 is imposed on the sample data i.e. no correlation. b) Numbers survived and fed over time for the true (blue) and empirical (red) estimates. An odds ratio of 100 is imposed on the sample data. c) An odds ratio of 0.0001 is imposed on the sample data.

## Results

The model predictions of the probability that mosquitoes are killed or alive and blood-fed in insecticide-treated experimental huts and how these effects change over time are presented in Figure 3 and 4. The median probable outcomes are reassuringly similar however the uncertainties for different models are very different.

Where only aggregated data are available, it is only possible to predict the alive and blood-fed probability if some assumption is made about independence between feeding and dying. Assuming independence using aggregated data predicts very similar median impacts to predictions made using data with the full complement of potential outcomes from a mosquito feeding attempt. However, behavioural vector biologists report that Anopheles gambiae sensu lato and *An. funestus* s.l. most often rest indoors after blood-feeding^27–33^, which indicates that blood-fed mosquitoes are possibly more likely to be exposed to insecticides sprayed onto indoor surfaces.

Only 3 datasets with a full breakdown of potential outcomes were available (Table 1). Two studies were conducted in Benin and 1 study was completed in Côte d’Ivoire, although both tested *An gambiae* s.l. mosquitoes in West African huts with both walls and ceilings sprayed. The key difference was that the Côte d’Ivoire study provided holed nets for the study participants to sleep under during the trial whereas the Benin studies provided no netting for sleepers. We could observe that the data from Côte d’Ivoire killed mosquitoes for a shorter period than those from Benin (Figure 3). It is possible that these untreated bed nets provided an alternative surface for blood fed mosquitoes to rest upon and digest feeds. Almost all mosquitoes blood-feed throughout the studies (median estimate across studies and months since spraying; 91.5%, 95%CI; 68.4% - 100% mosquitoes are blood-fed).

We can see in Figure 5 that the difference between the different predictions can be significant over longer periods of time. The model using comprehensive data only is very uncertain. However, these data provide worthwhile additional information when combined with the aggregate data such that the proportion survived and fed at 12 months could be approximately 10% higher than the aggregate data only model.

A major advantage of the Bayesian evidence synthesis over the alternative methods is seen by the tightened uncertainty bounds predicted (see Figure 5). This is driven by the additional data available for this statistical framework. To understand how this prediction is affected by correlated data sources we explore the correlation coefficient that is bounded by some function of the marginal probabilities which makes using a straightforward multivariate normal sampler fragile. Instead, we use the odds ratio (OR) (which is unrestricted) to specify the association between the two binary variables and then convert this OR, for given marginal probabilities of the events under consideration, into a (valid) correlation rho to be used in multivariate binomial sampling. We can see from Figure 5a-c that when mortality and blood-feeding are positively correlated, the number of successfully fed mosquitoes is under-estimated. This is our data situation. Alternatively, when mortality and blood-feeding are negatively correlated the number of successfully fed is over-estimated. Increasing data in the comprehensive resource (ILD) will always be beneficial to the accuracy of the predictions.

## Discussion

Concerns around mosquito resistance to pyrethroid insecticides has accelerated development of alternative insecticide products to help mitigate against any diminishing impact from pyrethroid-based vector control. By necessity, these products have different mechanisms of action so that rigorous testing is required prior to determining their potential benefit. This generates a wealth of data. Alongside this, statistical methods are developing and their availability to biological scientists is increasing through software languages such as R, BUGS and Stan. We demonstrate with these data how statistical models can be adapted and developed to maximise performance of inference from different but related data sources. This is all the more important since there are ethical decisions to be made when conducting such trials that result in differences. A key example is the debate about providing an untreated (sometimes holed) net for the study participant present to lure mosquitoes indoors during an IRS hut study. This is necessary since providing an intact net during an IRS study will prevent most blood-feeding which will then affect mosquito behaviour as IRS is designed primarily to target mosquitoes that are resting on walls after blood-feeding. In an IRS hut without a net most mosquitoes will feed and then rest on walls to digest the blood meal and are then killed by the IRS. A further complication is that, if a mosquito encounters a net, she may alight on the wall unfed and be killed by the IRS in which case the proportion of mosquitoes that are unfed-dead will be higher, as will be the proportion unfed-live mosquitoes, which may go on to feed elsewhere and transmit malaria.

Bayesian evidence synthesis (Model 3) allows both the most available, aggregated data and most accurate, comprehensive data to inform predictions of probable outcomes of mosquito feeding attempts. We show that this approach can refine predictions and tighten uncertainty. In practical terms, this may significantly increase or decrease the predicted proportions and so potentially impact decision making. The volume of aggregated data relative to comprehensive data means that the inference is shrunk towards that of Models 1 and 2 (Figures 3, 4c and 4d) which use aggregated data alone. With a greater volume of comprehensive data, which would be the ideal, these predictions might be altered. Simulating a correlated Bernoulli random variable allows us to understand how these biases play out; essentially, where probabilities are positively correlated (in our case, the probability that mosquitoes will be killed is correlated with the probability that mosquitoes will also be fed), we would expect the evidence synthesis to under-estimate the probability of mosquitoes blood-feeding and surviving. Conversely, were these probabilities negatively correlated then those mosquitoes surviving having blood-fed would be overestimated. In the context of IRS impact on mosquito behaviour, both scenarios are problematic. The first would overestimate the impact of the intervention while the second would undersell its potential. Acknowledging these effects of the statistical approach are therefore crucial.

We recognise that other, related modelling choices were available. For example, in terms of data, we could have aggregated the comprehensive data and combined these with the originally aggregated data to create a new aggregated data set. In terms of modelling, the aggregate data-only analysis assumed independent Binomial models for death and successfully fed. We could have estimated the two outcomes jointly as in the full Bayesian evidence synthesis model. However, the aim of this work was to compare approaches performed previously^16,17,26^ against the full Bayesian evidence synthesis approach, which we claim is the best option.

Alternative approaches to evidence synthesis are published which could be explored for their suitability to the peculiarities of the example used here, such as partial reconstruction of comprehensive data^34^. Another alternative is to include an indicator to the linear predictor to note those studies with additional data or potentially those where some assumption like independence is made a *priori* and allow them to have more weight in the ultimate predictions^35^. There are also sources of uncertainty in the data that we do not explore here including differences in wall surface, the provision of, and number of holes in, untreated mosquito nets for volunteer sleepers, the geographic location of the hut trial and respective mosquito species represented. Other studies have begun to consider the scale and implications of these differences^36–38^.

Given the increased capacity to collate multiple data and the increased access to statistical software, it is increasingly important to explore how different methods compare and infer biologically relevant conclusions. Bayesian evidence synthesis provides a robust statistical approach when different resources are available but may be biased toward the dataset with the greatest absolute quantity of information.

## Data Availability

The data is available within the paper.

## Supplementary Material

Supplementary Material 1. Modelling details, including BUGS and Stan code

Supplementary Material 2. Correlated binomial simulation

Supplementary data file 1. Datasets for analyses

## References

1. Bhatt S, Weiss DJ, Cameron E, et al. The effect of malaria control on Plasmodium falciparum in Africa between 2000 and 2015. Nature [Internet]. Nature Publishing Group, a division of Macmillan Publishers Limited. All Rights Reserved.; 2015 Oct 8 [cited 2017 Jan 12];526(7572):207–11. Available from: http://dx.doi.org/10.1038/nature15535

2. World Health Organization. Guidelines for malaria vector control [Internet]. Geneva; 2019. Available from: https://apps.who.int/iris/bitstream/handle/10665/310862/9789241550499-eng.pdf?ua=1

3. Sherrard-Smith E, Griffin JT, Winskill P, et al. Systematic review of indoor residual spray efficacy and effectiveness against Plasmodium falciparum in Africa. Nat Commun [Internet]. Nature Publishing Group; 2018 Dec 26 [cited 2018 Nov 27];9(1):4982. Available from: http://www.nature.com/articles/s41467-018-07357-w

4. Pluess B, Tanser FC, Lengeler C, Sharp BL. Indoor residual spraying for preventing malaria. Cochrane Database Syst Rev. Wiley; 2010 Apr 14;

5. Choi L, Pryce J, Garner P. Indoor residual spraying for preventing malaria in communities using insecticide-treated nets. Cochrane Database Syst Rev [Internet]. 2019 May 23 [cited 2019 Dec 20]; Available from: http://doi.wiley.com/10.1002/14651858.CD012688.pub2

6. Riley RD, Steyerberg EW. Meta-analysis of a binary outcome using individual participant data and aggregate data. Res Synth Methods. 2010 Jan;1(1):2–19.

7. Walraven C van. Individual patient meta-analysis—rewards and challenges. J Clin Epidemiol [Internet]. Pergamon; 2010 Mar 1 [cited 2019 Jun 4];63(3):235–237. Available from: https://www.sciencedirect.com/science/article/pii/S0895435609001103?via%3Dihub

8. Debray TPA, Moons KGM, Valkenhoef G van, et al. Get real in individual participant data (IPD) meta-analysis: a review of the methodology. Res Synth Methods [Internet]. John Wiley & Sons, Ltd; 2015 Dec 1 [cited 2019 Jun 4];6(4):293–309. Available from: http://doi.wiley.com/10.1002/jrsm.1160

9. Berlin JA, Santanna J, Schmid CH, Szczech LA, Feldman HI. Individual patient-versus group-level data meta-regressions for the investigation of treatment effect modifiers: ecological bias rears its ugly head. Stat Med [Internet]. John Wiley & Sons, Ltd; 2002 Feb 15 [cited 2019 Jun 4];21(3):371–387. Available from: http://doi.wiley.com/10.1002/sim.1023

10. Lambert PC, Sutton AJ, Abrams KR, Jones DR. A comparison of summary patient-level covariates in meta-regression with individual patient data meta-analysis. J Clin Epidemiol [Internet]. Pergamon; 2002 Jan 1 [cited 2019 Jun 4];55(1):86–94. Available from: https://www.sciencedirect.com/science/article/pii/S0895435601004140

11. Debray TPA, Damen Jaag, Snell KIE, et al. A guide to systematic review and meta-analysis of prediction model performance. BMJ. 2017 Jan;i6460.

12. Mai Y, Zhang Z. Software Packages for Bayesian Multilevel Modeling. Struct Equ Model A Multidiscip J [Internet]. Routledge; 2018 Jul 4 [cited 2019 Jun 4];25(4):650–658. Available from: https://www.tandfonline.com/doi/full/10.1080/10705511.2018.1431545

13. Brimacombe M. Likelihood methods in biology and ecology[]: a modern approach to statistics [Internet]. 1st Editio. Chapman and Hall CRC Press Taylor & Francis Group; 2019 [cited 2019 Jun 4]. Available from: https://books.google.co.uk/books?hl=en&lr=&id=9MuCDwAAQBAJ&oi=fnd&pg=PP1&dq=bayesian+statistical+modelling+for+ecologists+Stan+and+WinBugs&ots=F8-JuJnsQS&sig=mT94cV9QpmgSyqrB5rVNP613wXw#v=onepage&q&f=false

14. Riley RD, Moons KGM, Snell KIE, et al. A guide to systematic review and meta-analysis of prognostic factor studies. BMJ [Internet]. British Medical Journal Publishing Group; 2019 Jan 30 [cited 2019 Jun 4];364:k4597. Available from: http://www.ncbi.nlm.nih.gov/pubmed/30700442

15. Jackson C, Presanis A, Conti S, Angelis D De. Value of Information: Sensitivity Analysis and Research Design in Bayesian Evidence Synthesis. J Am Stat Assoc [Internet]. Taylor & Francis; 2019;114(528):1436–1449. Available from: https://doi.org/10.1080/01621459.2018.1562932

16. Mcdonald SA, Presanis AM, Angelis D De, Hoek W Van Der, Hooiveld M. An evidence synthesis approach to estimating the incidence of seasonal influenza in the Netherlands. 2013;33–41.

17. Jackson D. Confidence intervals for the between-study variance in random effects meta-analysis using generalised Cochran heterogeneity statistics. Res Synth Methods [Internet]. John Wiley & Sons, Ltd; 2013 Jun 1 [cited 2019 Jun 19];4(3):220–229. Available from: http://doi.wiley.com/10.1002/jrsm.1081

18. World Health Organization. Report of the Sixteenth WHOPES Working Group Meeting [Internet]. Geneva, Switzerland; 2013. Available from: http://www.who.int/whopes/en

19. Tchicaya ES, Nsanzabana C, Smith T a, et al. Micro-encapsulated pirimiphos-methyl shows high insecticidal efficacy and long residual activity against pyrethroid-resistant malaria vectors in central Côte d’Ivoire. Malar J [Internet]. 2014;13:332. Available from: http://www.pubmedcentral.nih.gov/articlerender.fcgi?artid=4159530&tool=pmcentrez&rendertype=abstract

20. Rowland M, Boko P, Odjo A, Asidi A, Akogbeto M, N’Guessan R. A new long-lasting indoor residual formulation of the organophosphate insecticide pirimiphos methyl for prolonged control of pyrethroid-resistant mosquitoes: an experimental hut trial in Benin. PLoS One [Internet]. 2013;8(7):e69516. Available from: http://journals.plos.org/plosone/article?id=10.1371/journal.pone.0069516

21. Agossa FR, Aikpon R, Azondekon R, et al. Efficacy of various insecticides recommended for indoor residual spraying: pirimiphos methyl, potential alternative to bendiocarb for pyrethroid resistance management in Benin, West Africa. Trans R Soc Trop Med Hyg [Internet]. Oxford University Press; 2014 Feb 1 [cited 2018 Jul 24];108(2):84–91. Available from: https://academic.oup.com/trstmh/article-lookup/doi/10.1093/trstmh/trt117

22. Oxborough RM, Kitau J, Jones R, et al. Long-lasting control of Anopheles arabiensis by a single spray application of micro-encapsulated pirimiphos-methyl (Actellic® 300 CS). Malar J [Internet]. BioMed Central; 2014 Jan 29 [cited 2018 Jul 24];13(1):37. Available from: http://malariajournal.biomedcentral.com/articles/10.1186/1475-2875-13-37

23. Lunn D, Spiegelhalter D, Thomas A, Best N. The BUGS project: Evolution, critique and future directions. Stat Med [Internet]. John Wiley & Sons, Ltd; 2009 Nov 10 [cited 2019 Jun 17];28(25):3049–3067. Available from: http://doi.wiley.com/10.1002/sim.3680

24. R Core Team. R: A Language and Environment for Statistical Computing [Internet]. Vienna, Austria: R Foundation for Statistical Computing; 2018. Available from: https://www.r-project.org/

25. Stan Development Team. Stan modeling language user’s guide and reference manual. [Internet]. Stan Development Team; 2017. p. 625. Available from: http://mc-stan.org/

26. Riley RD, Simmonds MC, Look MP. Evidence synthesis combining individual patient data and aggregate data: a systematic review identified current practice and possible methods. J Clin Epidemiol. 2007;60(5):431.e1-431.e12.

27. Paaijmans KP, Thomas MB. The influence of mosquito resting behaviour and associated microclimate for malaria risk. Malar J [Internet]. BioMed Central; 2011 Dec 7 [cited 2019 Jun 19];10(1):183. Available from: https://malariajournal.biomedcentral.com/articles/10.1186/1475-2875-10-183

28. Highton RB, Bryan JH, Boreham PFL, Chandler JA. Studies on the sibling species Anopheles gambiae Giles and Anopheles arabiensis Patton (Diptera: Culicidae) in the Kisumu area, Kenya. Bull Entomol Res [Internet]. Cambridge University Press; 1979 Apr 10 [cited 2019 Jun 19];69(1):43–53. Available from: https://www.cambridge.org/core/product/identifier/S0007485300017879/type/journal_article

29. Mnzava AE, Rwegoshora RT, Wilkes TJ, Tanner M, Curtis CF. Anopheles arabiensis and An. gambiae chromosomal inversion polymorphism, feeding and resting behaviour in relation to insecticide house-spraying in Tanzania. Med Vet Entomol [Internet]. 1995 Jul [cited 2019 Jun 19];9(3):316–24. Available from: http://www.ncbi.nlm.nih.gov/pubmed/7548951

30. Githeko AK, Service MW, Mbogo CM, Atieli FK. Resting behaviour, ecology and genetics of malaria vectors in large scale agricultural areas of Western Kenya. Parassitologia [Internet]. 1996 Dec [cited 2019 Jun 19];38(3):481–9. Available from: http://www.ncbi.nlm.nih.gov/pubmed/9257337

31. Faye O, Konate L, Mouchet J, et al. Indoor Resting by Outdoor Biting Females of Anopheles gambiae Complex (Diptera: Culicidae) in the Sahel of Northern Senegal. J Med Entomol [Internet]. 1997 May 1 [cited 2019 Jun 19];34(3):285–289. Available from: http://www.ncbi.nlm.nih.gov/pubmed/9151491

32. Rajaonarivelo V, Goff G Le, Cot M, Brutus L. Les anophèles et la transmission du paludisme à Ambohimena, village de la marge occidentale des Hautes-Terres Malgaches. Parasite [Internet]. 2004 Mar 25 [cited 2019 Jun 19];11(1):75–82. Available from: http://www.ncbi.nlm.nih.gov/pubmed/15071831

33. Killeen GF. Characterizing, controlling and eliminating residual malaria transmission. Malar J [Internet]. BioMed Central; 2014 Aug 23 [cited 2018 May 3];13(1):330. Available from: http://malariajournal.biomedcentral.com/articles/10.1186/1475-2875-13-330

34. Turner RM, Omar RZ, Yang M, Goldstein H, Thompson SG. A multilevel model framework for meta-analysis of clinical trials with binary outcomes. Stat Med [Internet]. 2000 Dec 30 [cited 2019 Jun 17];19(24):3417–3432. Available from: http://doi.wiley.com/10.1002/1097-0258%2820001230%2919%3A24%3C3417%3A%3AAID-SIM614%3E3.0.CO%3B2-L

35. Turner RM, Spiegelhalter DJ, Smith GCS, Thompson SG. Bias modelling in evidence synthesis. J R Stat Soc Ser A Stat Soc [Internet]. Wiley-Blackwell; 2009 Jan [cited 2019 Jun 17];172(1):21–47. Available from: http://www.ncbi.nlm.nih.gov/pubmed/19381328

36. Oumbouke WA, Fongnikin A, Soukou KB, Moore SJ, N’Guessan R. Relative performance of indoor vector control interventions in the Ifakara and the West African experimental huts. Parasit Vectors [Internet]. Parasites & Vectors; 2017;10(1):432. Available from: http://www.ncbi.nlm.nih.gov/pubmed/28927465 http://www.pubmedcentral.nih.gov/articlerender.fcgi?artid=PMC5606011

37. Massue DJ, Kisinza WN, Malongo BB, et al. Comparative performance of three experimental hut designs for measuring malaria vector responses to insecticides in Tanzania. Malar J [Internet]. BioMed Central; 2016;15(1):165. Available from: http://www.pubmedcentral.nih.gov/articlerender.fcgi?artid=4793500&tool=pmcentrez&rendertype=abstract

38. Briët OJ, Smith TA, Chitnis N. Measurement of overall insecticidal effects in experimental hut trials. Parasit Vectors [Internet]. BioMed Central; 2012 Nov 13 [cited 2017 Dec 13];5(1):256. Available from: http://parasitesandvectors.biomedcentral.com/articles/10.1186/1756-3305-5-256

